# Sex Differences in the Comorbidity between Attention Deficit-Hyperactivity Disorder and Posttraumatic Stress Disorder: A Systematic Literature Review and Meta-Analysis

**DOI:** 10.1101/2025.01.10.25320323

**Authors:** Julia Wilson, Doruntina Fida, Rie Maurer, Aleta Wiley, Therese Rajasekera, Primavera Spagnolo

## Abstract

**Objective:** Attention Deficit-Hyperactivity Disorder (ADHD) and Posttraumatic Stress Disorder (PTSD) are often comorbid and share a common core of symptoms. However, sex and gender-related factors significantly influence their prevalence, clinical presentation, and diagnosis. Here, we conducted a systematic literature review and meta-analysis to examine sex differences in ADHD/PTSD comorbidity during childhood and adulthood.

**Methods:** A scoping review of PsycINFO and PubMed yielded 13 eligible studies with complete outcome data. We conducted fixed-effects meta-analyses of the sex-stratified prevalence of ADHD/PTSD using pooled odds ratios (OR) with a 95% confidence interval (CI). Fixed-effects subgroup analyses were performed using age as a subgroup. Effect size heterogeneity was assessed using the I^2^ index and Cochran’s Q test.

**Results:** In the whole sample (N= 13,585; F= 7005, M= 6580), the diagnosis of ADHD/PTSD was significantly higher in females than in males (OR = 1.32, *p* = 0.02). Between-study heterogeneity was low-to-moderate and not significant (I^2^ = 41%; *p* = 0.06), validating the fixed-effects model. Age-stratified subgroup analyses revealed higher ADHD/PTSD odds in females compared to males only in adult populations (OR=1.41; *p =* 0.01). Additionally, females were more likely to be diagnosed with both disorders in studies where ADHD was the primary diagnosis (OR = 1.60; *p =* 0.002), and in studies employing structured clinical interviews as diagnostic tools (OR = 1.46; *p =* 0.009).

**Conclusions:** Our study is the first to show that the association between ADHD and PTSD is stronger in females, suggesting that ADHD may increase risk for PTSD in a sex-specific manner.

## Introduction

Attention Deficit-Hyperactivity Disorder (ADHD) and Posttraumatic Stress Disorder (PTSD) are two of the most common psychiatric disorders in the United States, with estimated lifetime prevalence rates of 8.2% and 6.8%, respectively [1]. Though clinically distinct, ADHD and PTSD often co-occur, with comorbidity rates ranging between 2% to 37% across the lifespan [2, 3]. These disorders also exhibit a common core of behavioral and cognitive manifestations, including attentional deficits, dysregulated affect, hyperarousal, and hypervigilance [4, 5, 6], and share similar neural alterations, particularly an abnormal activation of brain regions implicated in fear processing [7, 8]. Moreover, individuals with comorbid ADHD and PTSD exhibit more severe symptomatology for both disorders [9, 10] and a higher prevalence of other psychiatric comorbidities compared to subjects with ADHD or PTSD alone [11]. The significant association between PTSD and ADHD suggests that these disorders might be causally linked, as indicated by both retrospective and longitudinal studies showing that ADHD represents an important risk factor for PTSD [8, 10, 11, 12].

However, these disorders also exhibit important differences in their prevalence and clinical presentation, which appear to be driven by the effects of both biological sex and gender-related factors. Specifically, epidemiological studies indicate that ADHD is male-predominant disorder, with a female-to-male ratio of 1:4 in children, which becomes closer to 1:2 in adults [13, 14, 15, 16]. Conversely, PTSD is more common in females, with a female-to-male ratio ranging from 2:1 to 3:1, in both children and adults. Sex differences in type and severity of symptoms as well as in treatment response have also been reported for both ADHD and PTSD [17, 18, 19, 20]. These differences may be partly explained by gender-related factors, such as under-recognition of ADHD in girls [18, 21, 22] and greater rates of exposure to high-impact trauma (e.g., sexual trauma) in women compared to men [17].

Sex-specific factors also seem to be associated with quantitative and qualitative differences in ADHD and PTSD between individuals of different birth-assigned sexes. For instance, carriers of sex chromosome aneuploidies have an increased risk of ADHD symptoms, particularly in association with male phenotypes [23, 24, 25]. Steroid hormones, particularly estradiol, exert a larger influence on mood in females with ADHD compared with those without ADHD [26] and play a major role in modulating fear response mechanisms involved in PTSD in females [27, 28, 29, 30].

Despite this body of evidence, no study has specifically examined whether the association between ADHD and PTSD varies between sexes. To begin addressing this knowledge gap, we conducted a systematic review and meta-analysis of the extant literature on the relationship between ADHD and PTSD to uncover sex differences in ADHD/PTSD comorbidity during childhood and adulthood. We hypothesized that the odds of ADHD and PTSD comorbidity would vary by sex, with women showing a stronger association compared to men.

## Methods

### Data Sources

This study was conducted in line with the Preferred Reporting Items for Systematic Reviews and Meta-Analyzes (PRISMA) 2020 Statement Guidelines [31] and was registered on PROSPERO (ID: CRD42020179855). We performed a systematic literature search of all the journal articles available through PubMed and PsycINFO published between January 1, 2000, and October 23, 2023, as well as the reference lists of eligible studies. The search strings used were (“PTSD” OR “Post-Traumatic Stress Disorder” OR “Posttraumatic Stress Disorder”) AND (“ADHD” OR “attention deficit hyperactivity disorder” OR “ADD” or “attention deficit disorder”). Additionally, reference lists of included articles were screened to identify further articles for inclusion. We excluded reviews and articles not written in English, and studies evaluating subjects exclusively for PTSD or ADHD. After duplicates were removed, the search yielded 74 articles, which were screened for inclusion by three authors. Disagreements among the authors were resolved by the senior author.

### Study Selection

Studies were included in the meta-analysis if they: i) represented original human subjects research, ii) contained a comparison or control group, iii) reported numbers of participants with diagnoses of ADHD and PTSD, iv) diagnosed ADHD and PTSD using validated diagnostic tools, v) compared rates of PTSD in ADHD subjects vs controls, or rates of ADHD in PTSD subjects vs controls; vi) included sex-stratified prevalence rates of ADHD and PTSD. Excluded articles were studies that failed to meet any one of the above criteria. Reviews, letters, and editorials were also excluded.

### Data Extraction and Analysis

One author (JW) independently reviewed all primary studies. A second author (DF) checked any data discrepancies, and any disagreements between authors were resolved by the senior author (PAS). The following information was extracted from all included articles: 1) authors; 2) year of publication; 3) title; 4) sample size; 5) age (mean or median and SD) and age group (*pediatric* or *adult*); 6) prevalence of ADHD in PTSD and controls OR prevalence of PTSD in ADHD and controls, overall and by sex; 7) number and sex distribution of subjects with and without ADHD and PTSD alone and combined; 8) methods of PTSD and ADHD diagnostic assessment; 9) type of control group (healthy or psychiatric control group). When available, we also extracted the race/ethnicity of subjects included in the studies. For studies that provided total N and percentages without numbers of cases, we used the percentages provided to calculate raw frequencies. Furthermore, for studies that did not include sex-stratified data, attempts were made to contact the researchers to obtain such data.

Odds ratios (OR) were calculated with a 95% confidence interval (CI) for the raw frequencies of comorbid ADHD and PTSD, stratified by sex. A fixed-effects meta-analysis of the sex-stratified ADHD/PTSD prevalence was conducted, with male sex as the reference group (Mantel-Haenszel method). We ran fixed-effects subgroup analyses based on primary diagnosis (ADHD vs. PTSD), age (minors vs. adults), population sampled (clinical vs. community), and race (white vs non-white).

Odds ratios were computed using the {meta} package in RStudio and effect size heterogeneity was assessed using the I^2^ index. An Egger regression was performed to assess potential publication bias and visualized in a funnel plot (Figure 3). For all analyses, significance was determined if *p* < 0.05.

## Results

### Study selection and characteristics

A flowchart of the selection process is shown in Fig. 1. Our literature search produced 74 unique full-text articles, seven of which met the eligibility criteria. Six studies were further included after contacting the authors and obtaining sex-disaggregated data. Thus, our meta-analysis comprised a total of thirteen studies published between January 1, 2000 and October 23, 2023 (Table 1). Overall, these studies included a total of 13,858 subjects, with an approximate male-to-female ratio of 1:1 (6,580 males and 7,005 females). Seven studies were performed in adults (≥ 18 years old) and six in minors (< 18 years old). With regard to study design, 12 studies were cross-sectional [9, 11, 32, 33, 34, 35, 36, 37, 38, 39, 40, 41] and one was a longitudinal study [10]. As detailed in Table 1, the majority of studies examined the prevalence of PTSD in individuals with ADHD (n=8). Overall, diagnoses of both ADHD and PTSD were predominantly made using clinical interviews (11/13 studies). Only two studies reported data disaggregated by race [9, 10].

**Figure 1.**
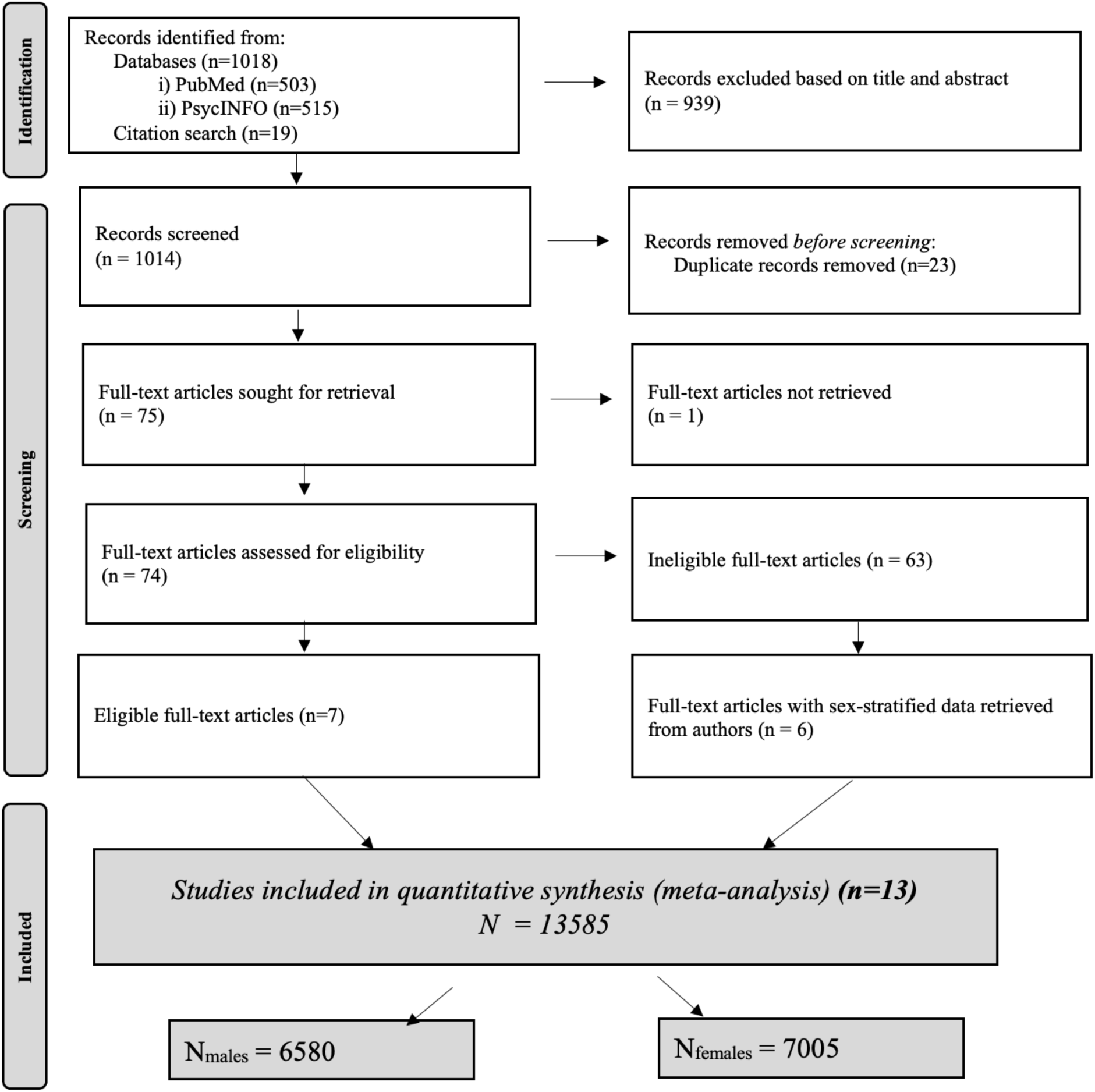
PRISMA Flow Diagram

**Table 1.**
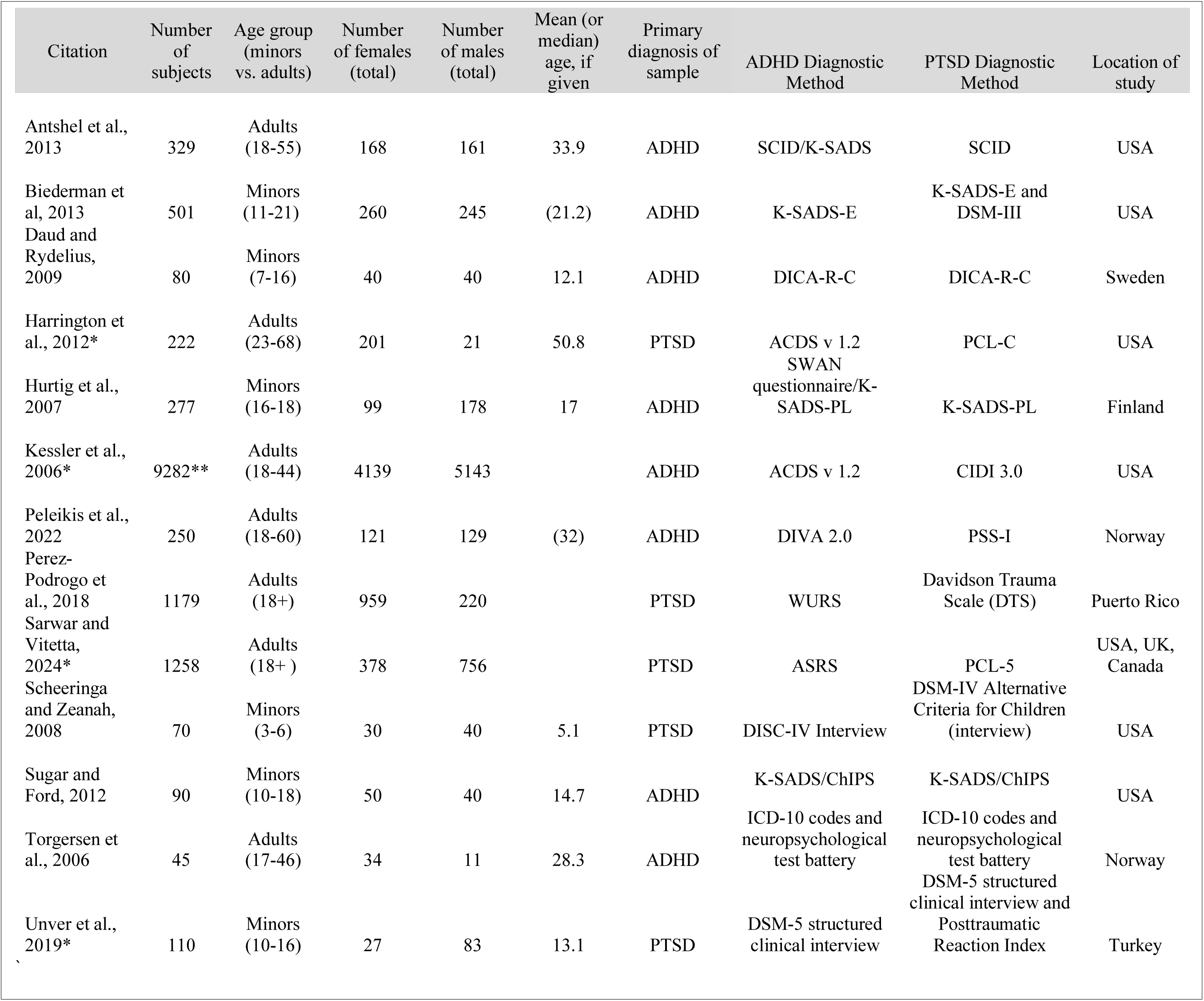
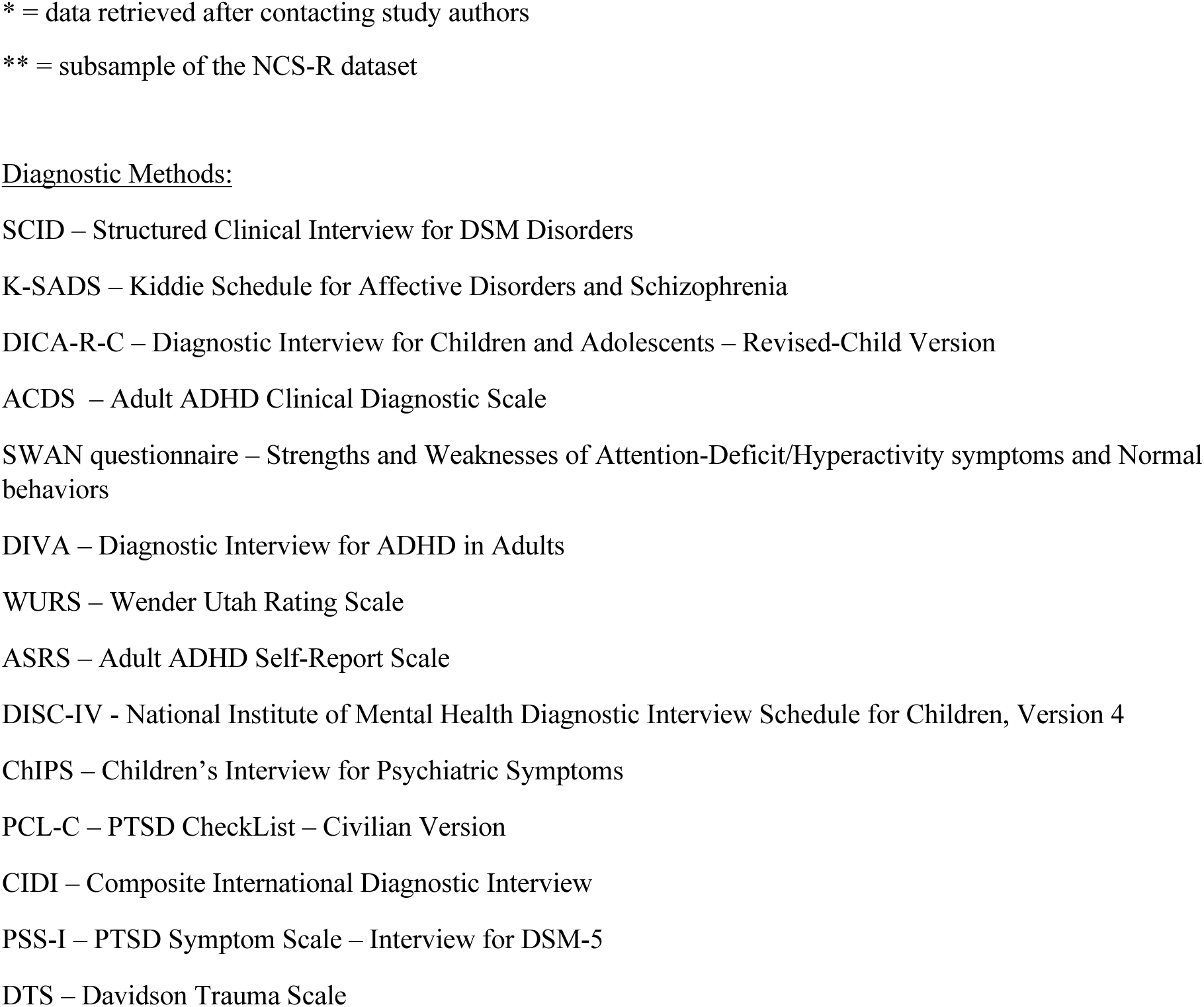
Studies Included in Systematic Review and Meta-Analysis.

### Meta-analytic results

A fixed-effects meta-analysis of the whole sample (n= 13,585) yielded 1.32 increased odds of comorbid ADHD/PTSD in females compared to males (95% CI [1.04; 1.66]; *p*= 0.02, Figure 1). When controls were excluded from the analysis, a similar trend was observed, although it was not significant (OR=1.12; 95% CI [0.90 - 1.47]; *p*= 0.26). Between-study heterogeneity was low-to-moderate (I^2^ = 41%; Figure 2) and not significant (*p*= 0.06), validating the fixed-effects model. The Egger regression testing for publication bias was also not significant (*p* = 0.81, see Figure 3).

**Figure 2.**
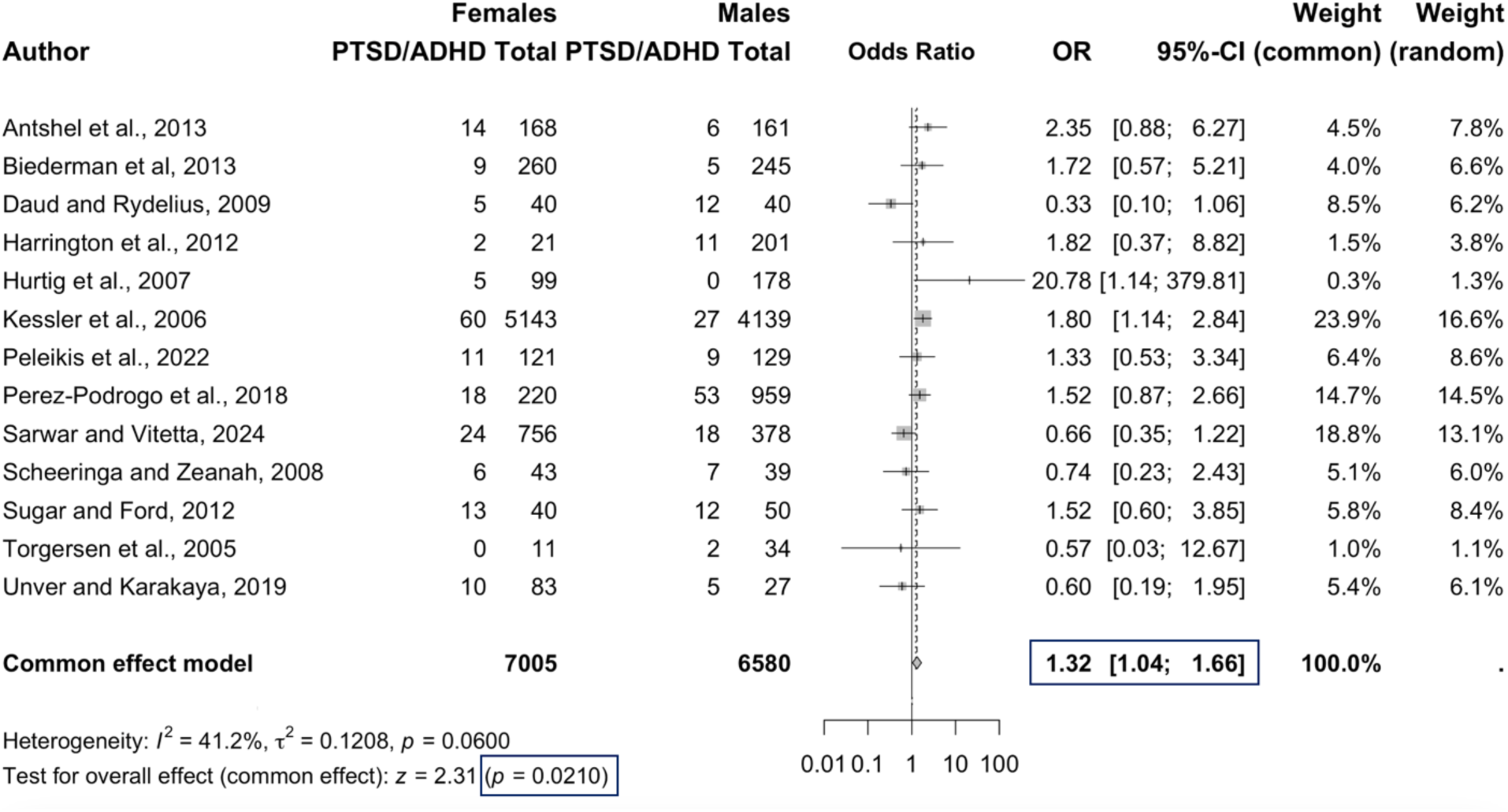
Odds Ratio of Comorbid PTSD/ADHD in Females Compared to Males

**Figure 3.**
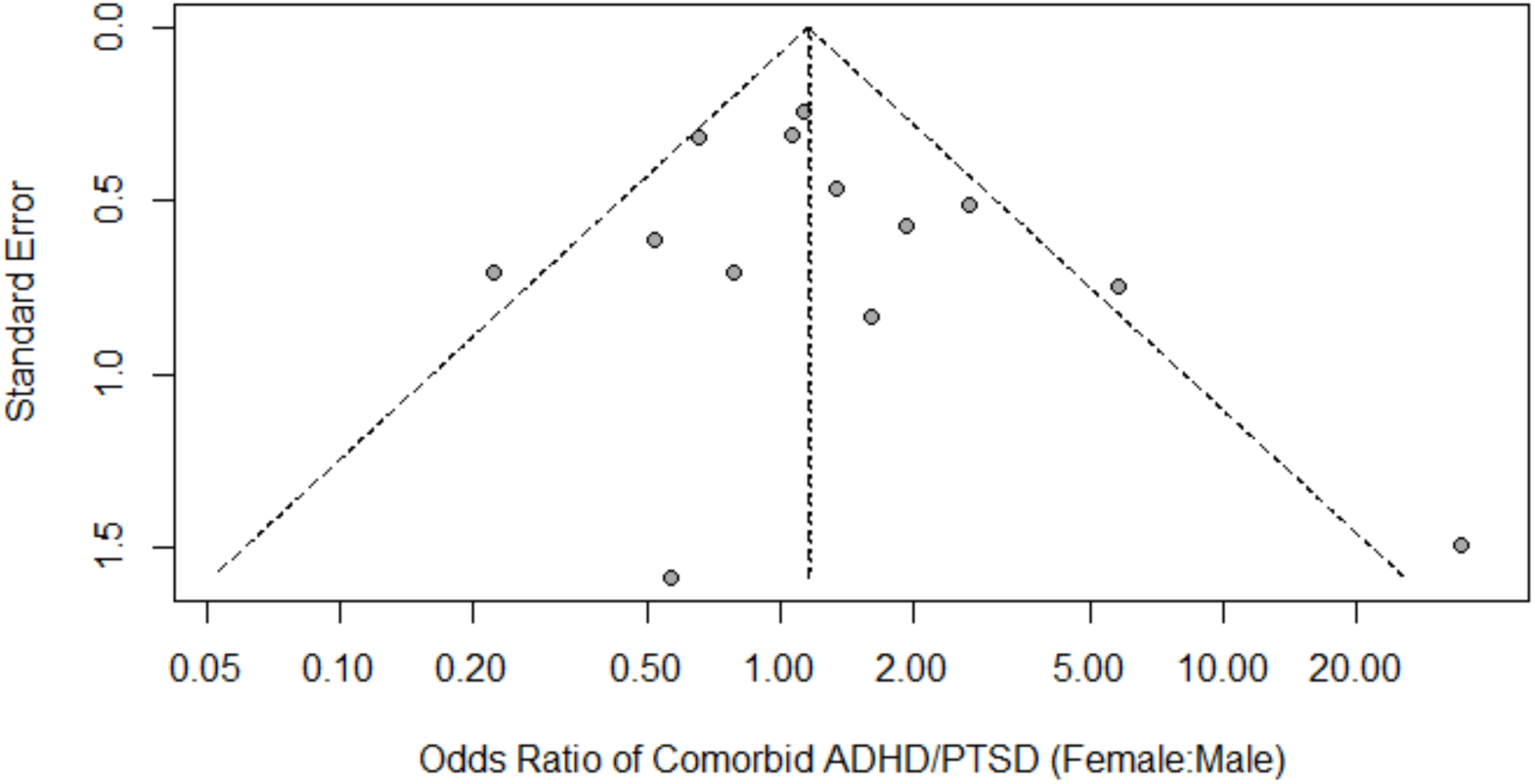
Funnel Plot of Between-Study Heterogeneity of Female: Male ADHD/PTSD Odds Ratio

Next, we computed a separate meta-analysis for studies examining ADHD in individuals with PTSD and vice versa and observed that the prevalence of comorbid ADHD/PTSD in females vs males significantly differed among these subgroups (*p=* 0.045). Specifically, we found that females were significantly more likely to be diagnosed with comorbid ADHD and PTSD than males in studies assessing the prevalence of PTSD in subjects with ADHD [8 studies [9, 10, 31, 32, 33, 34, 38, 40]; *n*= 10,858] (OR = 1.60; 95% CI [1.18 - 2.17]; *p =* 0.002), while no sex/gender differences were observed in studies examining rates of ADHD in subjects with PTSD [5 studies [11, 35, 36, 37, 39]; *n*=2,727] (OR = 0.98; 95% CI [0.67 - 1.42]; *p* = 0.91).

We further found that in adult studies [7 studies [9, 11, 34, 35, 36, 37, 41]; *n*= 12,441], the odds of having comorbid ADHD/PTSD were 1.41 times higher in women compared to men (OR=1.41; 95% CI [1.08 - 1.86]; *p =* 0.01), whereas they did not significantly differ among sexes in pediatric studies [6 studies [10, 32, 33, 38, 39, 40]; *n*= 1,144] (OR=1.08; 95% CI [0.67 - 1.70]; *p =* 0.75). Among both clinical (8 studies; *n*=1,631) and community samples (5 [33, 34, 36, 37, 38]; *n*=11,954), women represented the majority of individuals with comorbid ADHD/PTSD, although the OR was significantly higher only in females vs males from community samples compared to clinical samples ([OR = 1.39; CI [1.04; 1.86]; *p*= 0.03] *vs* [OR = 1.19, 95% CI [0.81 - 1.77]; *p* = 0.37]).

Among the 11 studies [9, 10, 11, 32, 33, 34, 35, 38, 39, 40, 41] that used a structured clinical interview to assess diagnoses of ADHD and PTSD (*n*=11,272), females had significantly increased odds of comorbid ADHD/PTSD (OR = 1.46; 95% CI [1.10 - 1.93]; *p =* 0.009). However, females and males were equally likely to be diagnosed with ADHD/PTSD among the two studies ([36, 37], *n*= 2,313) that employed rating scales (OR = 1.04; 95% CI [0.68 - 1.59]; *p* = 0.87). Between-study heterogeneity was not significant for any of the subgroups analyzed, indicating that fixed-effects analysis was appropriate for all subgroup meta-analyses performed (see Supplemental Figures 1-4).

Finally, we explored whether the odds of comorbid ADHD/PTSD varied according to the race of study participants. Only 2 studies (n = 833) reported rates of comorbid ADHD/PTSD by race [9, 10]. Both studies investigated the prevalence of PTSD in ADHD; one study was a longitudinal study in minors [10], while the other was a cross-sectional study in adults [9]. In these 2 samples, non-white individuals were significantly more likely to be diagnosed with both PTSD and ADHD than white individuals (OR = 2.39, CI [0.99 - 5.75], *p* = 0.045, Supplemental Figure 5). Given that none of the studies reported data disaggregated by both sex and race, we could not explore the effects of sex × race interaction on the association of ADHD and PTSD.

## Discussion

Building on the extensive evidence of a link between ADHD and PTSD [8, 42], the current study sought to systematically examine sex differences in the prevalence of ADHD/PTSD comorbidity and to uncover the influence of age, primary diagnosis, and diagnostic methods in modulating this relationship. In line with our hypothesis, we found that females had significantly increased odds of being diagnosed with comorbid ADHD and PTSD compared to males. Age-stratified subgroup analyses revealed that this association was significant in adult populations but not in pediatric populations. Additionally, females were more likely to be diagnosed with both disorders in studies where ADHD was the primary diagnosis and in studies employing structured clinical interviews as diagnostic tools. Conversely, the sampled population (clinical vs. community) did not modulate the sex-specific association between PTSD and ADHD. While the female-to-male odds ratio was only significantly increased in the community samples, this difference may be attributable to differences in sample size between the subgroups (*n* _clinical_ = 1,631; *n* _community_ = 11,954).

Our findings of a stronger association between ADHD and PTSD in females *vs* males, particularly during adulthood, expand on previous research showing that patterns of comorbidities in both PTSD and ADHD vary by sex. Furthermore, given that ADHD has been shown to increase risk for PTSD ([3, 12, 43]; but see also [44]), our results indicate that this disorder may represent a greater vulnerability factor for PTSD in women compared to men. At the behavioral level, ADHD is characterized by impulsivity and risk-taking [45], which may increase the risk for traumatic exposure, and consequently for PTSD [46]. Interestingly, several studies have indicated that sex biases in PTSD prevalence are at least partially explained by a greater exposure to high-impact trauma, and at a younger age, in women compared to men [17, 47, 48]. At the neurobiological level, ADHD has also been associated with alterations in the brain fear network, which regulates threat- and fear-related responses, thus playing a major role in the pathophysiology of PTSD [20, 48].

Accumulating evidence from studies employing fear conditioning/extinction paradigms shows significant sex differences in the activation and functional reactivity of the fear network, mainly due to the influence of sex hormones [28, 49, 50]. Specifically, decreased or fluctuating estrogen levels have been shown to reduce fear extinction and enhance return of fear, while high estrogen levels have the opposite effect, although they also seem to facilitate fear acquisition [28]. The complex and dynamic modulation that estrogen exerts on the neural machinery of fear processing across the female lifespan has been recognized as one of the key mechanisms underlying sex differences in PTSD vulnerability [17, 29]. Based on these observations, we may hypothesize that hormonal influences, which worsen ADHD severity in women, may also exacerbate the negative impact of ADHD-related brain abnormalities on the risk of developing PTSD. Future studies should examine whether fear network impairments in individuals with ADHD exhibit sex differences and/or modulate PTSD vulnerability in a sex-dependent manner.

We further observed that odds of ADHD/PTSD comorbidity were higher in females compared to males in studies assessing PTSD prevalence among individuals diagnosed with ADHD, whereas no sex differences were observed in studies where PTSD was the primary diagnosis. ADHD is frequently missed or diagnosed late in females, partly because of sex differences in disease presentation, with the male-predominant hyperactive-impulsive subtype being more easily identifiable compared to the inattentive subtype, which is more prevalent in females (for a review see [18]). In addition, the underdiagnosis of ADHD in females could also result from a primary recognition of co-occurring neuropsychiatric symptoms that overshadow underlying ADHD symptoms, or from an initial misdiagnosis with other mental health disorders [51, 52]. As such, our findings appear to further support the hypothesis that ADHD may represent a vulnerability factor for PTSD to which women are more susceptible than men. Indeed, we observed greater odds of ADHD/PTSD comorbidity in women with a primary diagnosis of ADHD, thus suggesting that gender biases in ADHD recognition did not influence the observed association. Our hypothesis is also in line with the results of a recent study showing that the impact of several risk factors for PTSD significantly differed among men and women [53].

It should, however, be noted that the relationship between ADHD and PTSD may be bidirectional [8]. This bidirectionality could be explained by shared genetic liability and common environmental risk factors, particularly childhood trauma, that may lead to brain endophenotypes (fear network impairments) conferring vulnerability to both conditions [12]. Further research is needed to understand whether these factors contribute to ADHD and PTSD risk in a sex-dependent manner and/or whether they differentially shape the directionality.

Lastly, we aimed to evaluate the intersectional effect of sex and race on the odds of comorbid ADHD/PTSD. However, none of the studies included in our meta-analysis reported data disaggregated by both sex and race. Given that no prior research had examined the influence of race on ADHD/PTSD comorbidity, we chose to leverage data from the only two studies reporting diagnosis rates by race [9, 10] to begin addressing this question. Our findings of a marginally stronger association of ADHD/PTSD comorbidity in racial minorities compared to white subjects warrant further investigation, since previous studies have also demonstrated increased prevalence of both disorders among racial minorities, particularly Black individuals [54, 55], who also seem to face greater disparities in receiving diagnosis [56, 57].

Our findings should be interpreted in light of several limitations. First, although our sample size was large, our analyses were limited to the studies meeting our eligibility criteria and reporting data disaggregated by sex. We hope that a broader application of federal legislation and national calls requesting authors to stratify and report outcome data by sex (and gender [see [58, 59]]) will facilitate future comparative and prospective studies. Furthermore, increased availability of sex-disaggregated data, such as age of onset and disease duration, may provide additional insights into the influence of sex and gender on the link between ADHD and PTSD. The methodological heterogeneity of the studies included in our meta-analysis represents a further limitation, which we partially addressed by conducting subgroup analyses based on sample characteristics and diagnostic methods.

These limitations notwithstanding, to the best of our knowledge, this systematic review and meta-analysis was the first to report that the association between ADHD and PTSD is stronger in women, suggesting that ADHD may increase risk for PTSD in a sex-specific manner. These findings support the need for more sex- and gender-informed investigation of factors shaping ADHD and PTSD risk, which can lead to the development of tailored prevention and treatment strategies.

## Data Availability

All data produced in the present study are available upon reasonable request to the authors.

https://www.doi.org/10.4088/JCP.12m07698

https://www.doi.org/10.1111/acps.12011

https://www.doi.org/10.1016/j.comppsych.2011.12.001

https://www.doi.org/10.1007/s00787-007-0607-2

https://www.doi.org/10.1176/ajp.2006.163.4.716

https://www.doi.org/10.1080/08039488.2021.1962973

https://www.doi.org/10.1016/j.psychres.2018.04.017

https://www.doi.org/10.4236/oalib.1111111

https://www.doi.org/10.1080/15374410802148178

https://www.doi.org/10.1002/jts.21668

## Acknowledgments

This work was conducted with support from the Mary Horrigan Connors Center for Women’s Health and Gender Biology and the Women’s Brain Health Initiative, Brigham and Women’s Hospital, Boston, MA, USA. We also acknowledge the support from UM1TR004408 award through Harvard Catalyst | The Harvard Clinical and Translational Science Center (National Center for Advancing Translational Sciences, National Institutes of Health) and financial contributions from Harvard University and its affiliated academic healthcare centers. The content is solely the responsibility of the authors and does not necessarily represent the official views of Harvard Catalyst, Harvard University and its affiliated academic healthcare centers, or the National Institutes of Health.

## Declaration of competing interest

The authors declare no competing interests.

## Notes

### Competing Interest Statement

The authors have declared no competing interest.

### Author Declarations

The study used (or will use) ONLY openly available human data that were originally located at: doi: 10.4088/JCP.12m07698 doi: 10.1111/acps.12011 doi: 10.1016/j.comppsych.2011.12.001 doi: 10.1007/s00787-007-0607-2 doi: 10.1176/ajp.2006.163.4.716 doi: 10.1080/08039488.2021.1962973 doi: 10.1016/j.psychres.2018.04.017 doi: 10.4236/oalib.1111111 doi: 10.1080/15374410802148178 doi: 10.1002/jts.21668 doi: 10.1080/08039480500520665 doi: 10.1177/1087054716677818

