## Supplementary figures and images for "Sex Differences in the Comorbidity between Attention Deficit-Hyperactivity Disorder and Posttraumatic Stress Disorder: A Systematic Literature Review and Meta-Analysis"

### Supplemental Figure 1

Supplemental Figure 1: ADHD/PTSD Subgroup Analysis by Primary Diagnosis
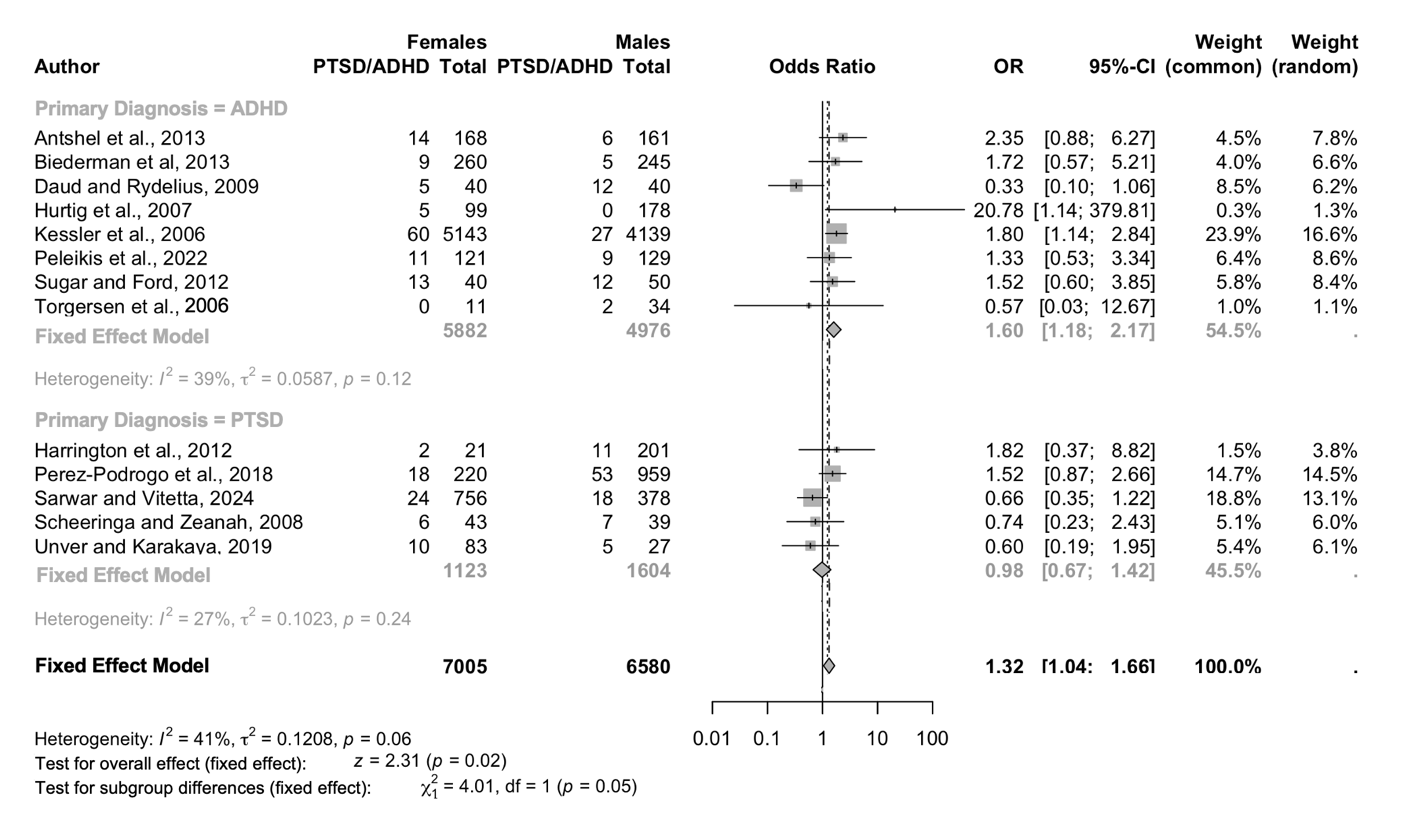

### Supplemental Figure 2

Supplemental Figure 2: ADHD/PTSD Subgroup Analysis by Age

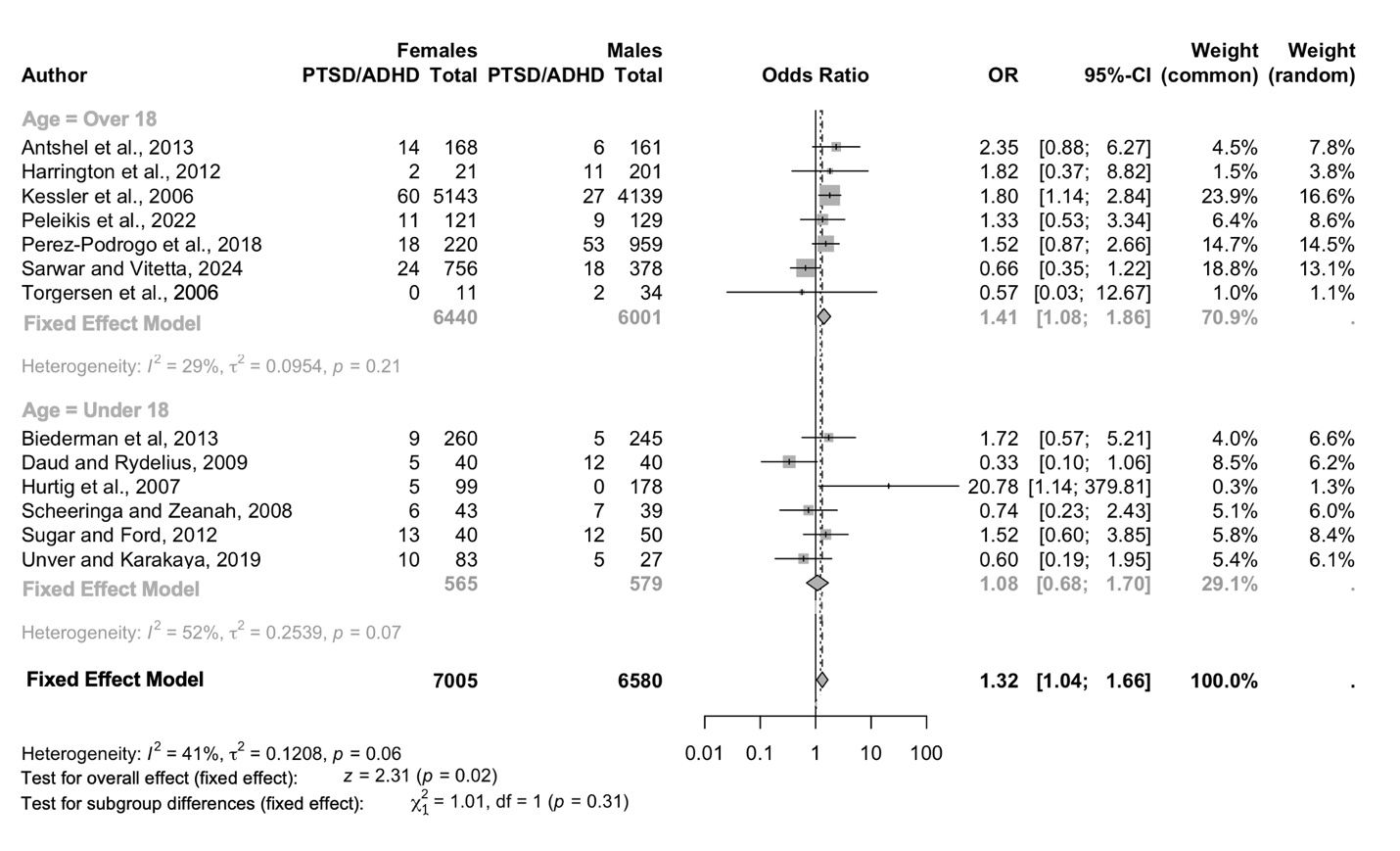

### Supplemental Figure 3

Supplemental Figure 3: ADHD/PTSD Subgroup Analysis by Subject Population

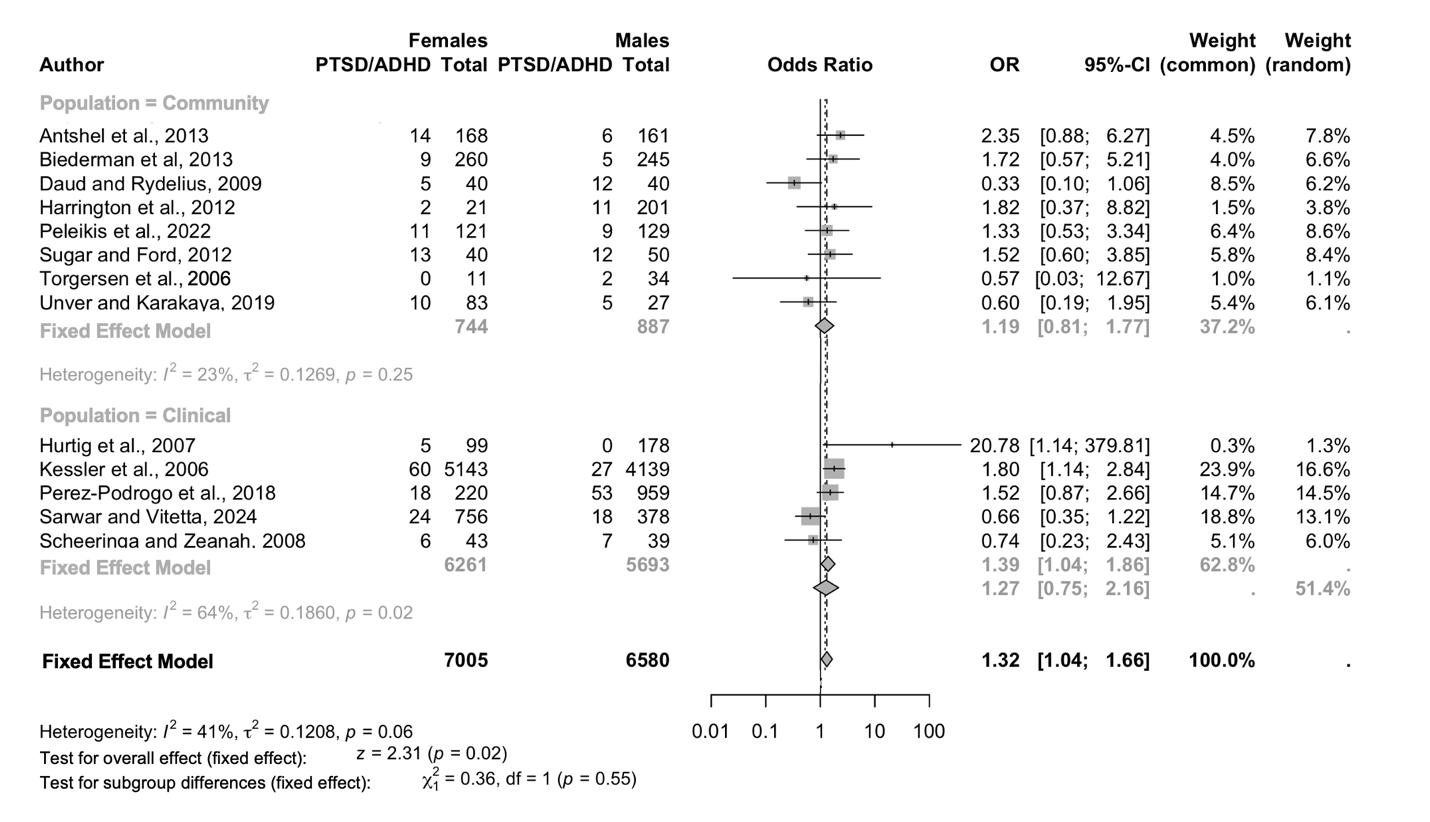

### Supplemental Figure 4

Supplemental Figure 4: ADHD/PTSD Subgroup Analysis by Diagnostic Method

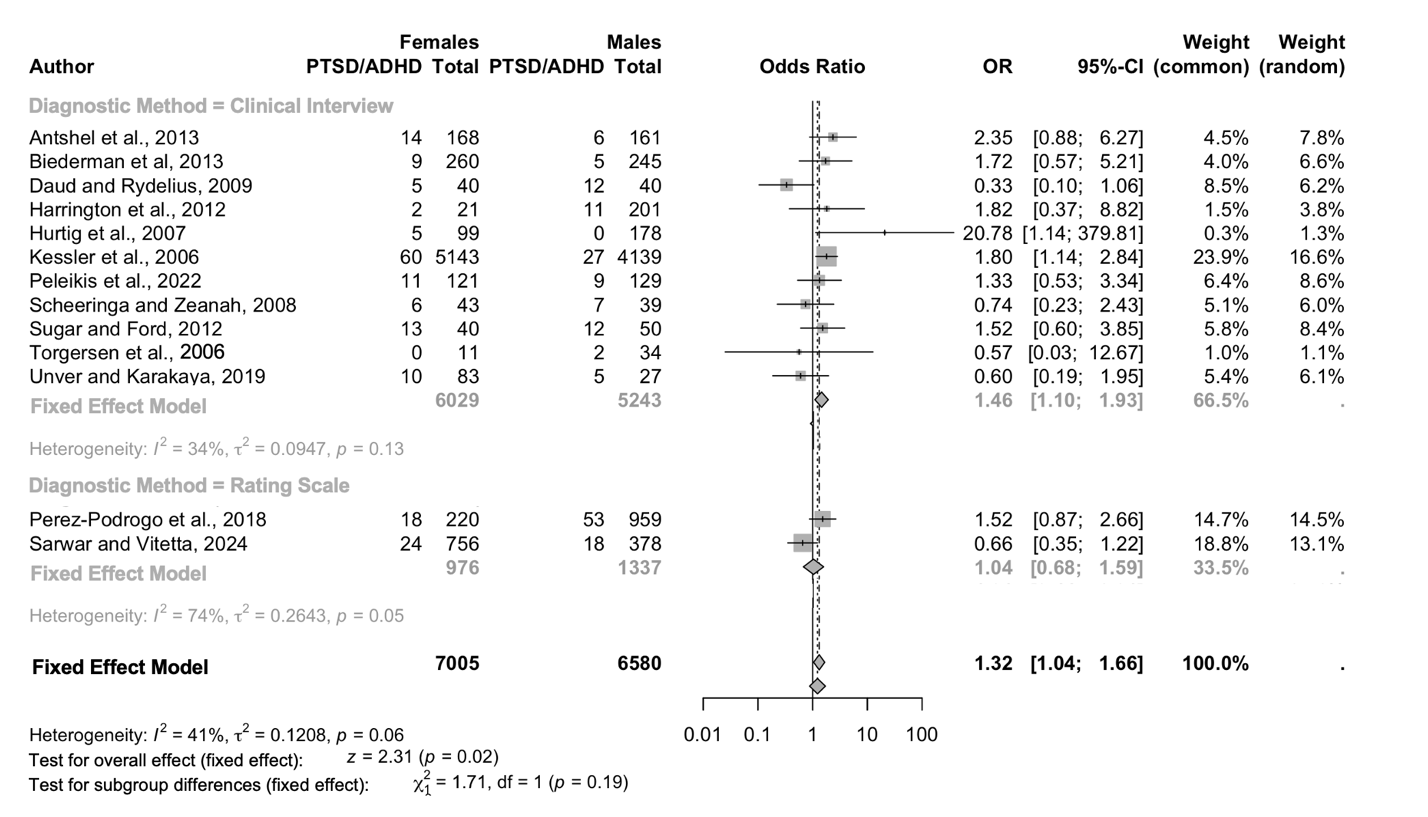

### Supplemental Figure 5

Supplemental Figure 5: ADHD/PTSD Odds Ratio by Race (Non-White vs. White)

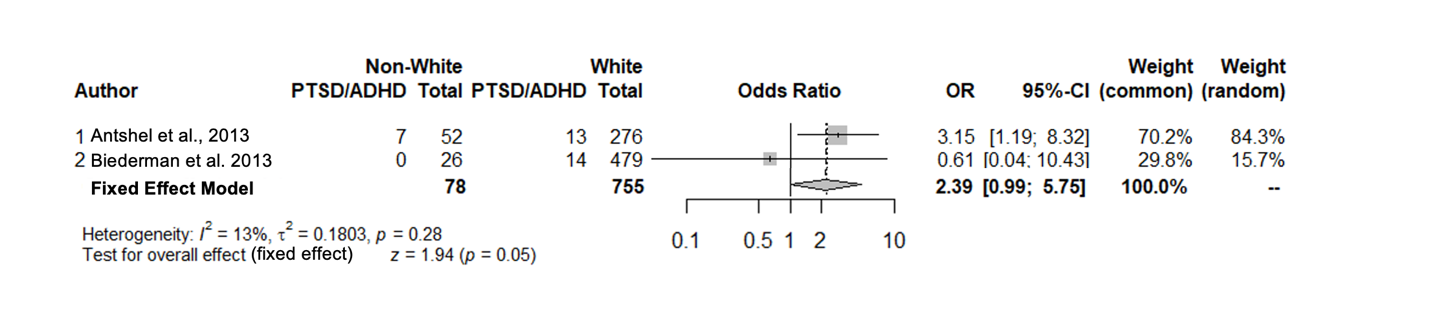
